# Validation of a Trans-Ancestry Polygenic Risk Score for Type 2 Diabetes in Diverse Populations

**DOI:** 10.1101/2021.09.11.21263413

**Authors:** Tian Ge, Amit Patki, Vinodh Srinivasasainagendra, Yen-Feng Lin, Marguerite Ryan Irvin, Hemant K. Tiwari, Nicole Armstrong, Brittney H. Davis, Emma Perez, Vivian Gainer, Barbara Benoit, Mark J. O’Connor, Renuka Narayan, Bethany Etheridge, Maria Stamou, Aaron Leong, Miriam S. Udler, Karmel W. Choi, Ayme D. Miles, Krzysztof Kiryluk, Atlas Khan, Chia-Yen Chen, Yen-Chen Anne Feng, Hailiang Huang, James J. Cimino, Shawn Murphy, Scott T. Weiss, Christoph Lange, Maggie C. Y. Ng, Jordan W. Smoller, Matthew S. Lebo, James B. Meigs, Nita A. Limdi, Elizabeth W. Karlson

## Abstract

Type 2 diabetes (T2D) is a worldwide scourge caused by both genetic and environmental risk factors that disproportionately afflicts communities of color. Leveraging existing large-scale genome-wide association studies (GWAS), polygenic risk scores (PRS) have shown promise to complement established clinical risk factors and intervention paradigms, and improve early diagnosis and prevention of T2D. However, to date, T2D PRS have been most widely developed and validated in individuals of European descent. Comprehensive assessment of T2D PRS in non-European populations is critical for an equitable deployment of PRS to clinical practice that benefits global populations. Here we integrate T2D GWAS in European, African American and East Asian populations to construct a trans-ancestry T2D PRS using a newly developed Bayesian polygenic modeling method, and evaluate the PRS in the multi-ethnic eMERGE study, four African American cohorts, and the Taiwan Biobank. The trans-ancestry PRS was significantly associated with T2D status across the ancestral groups examined, and the top 2% of the PRS distribution can identify individuals with an approximately 2.5-4.5 fold of increase in T2D risk, suggesting the potential of using the trans-ancestry PRS as a meaningful index of risk among diverse patients in clinical settings. Our efforts represent the first step towards the implementation of the T2D PRS into routine healthcare.

## INTRODUCTION

Type 2 diabetes (T2D) is a common, chronic disease caused by both genetic and environmental risk factors and their interactions (Dietrich et al. 2019), which has significantly increased prevalence in the past 20 years (Wang et al. 2021) and disproportionately afflicts African American and Latino communities (Gregg et al. 2013; Menke et al. 2015; Centers for Disease Control and Prevention 2020). The current screening of T2D focuses on individuals with demographic and clinical risk factors, including overweight or obesity, age >35 years, and a family history of diabetes (US Preventive Services Task Force 2021). However, despite preventative strategies and public health efforts to improve nutrition and physical activity, facilitate access to care, and limit tobacco and alcohol use, the morbidity and mortality associated with T2D remain unaltered (Centers for Disease Control and Prevention 2020), likely because most interventions are adopted too late in the course of disease trajectory.

Recent large-scale genome-wide association studies (GWAS) in diverse populations have identified hundreds of genetic loci associated with T2D (Mahajan et al. 2018; Mahajan et al. 2020; Vujkovic et al. 2020). Polygenic risk scores (PRS), which aggregate the genetic risk of individual alleles across the genome, are thus promising to predict future T2D occurrence and improve early diagnosis, intervention and prevention of T2D (Meigs et al. 2008; Hivert et al. 2011; Vassy et al. 2012a, 2012b, 2014;Walford et al. 2012). However, to date, T2D PRS were most widely developed and validated in individuals of European descent. Given that the predictive performance of PRS often attenuates in non-European populations (Martin et al. 2019), and communities of color are experiencing continuing increased rates of T2D (Gregg et al. 2013; Menke et al. 2015; Centers for Disease Control and Prevention 2020; Wang et al. 2021), it is critically important to assess and optimize the transferability of T2D PRS in diverse populations before they can be deployed in clinical settings.

The Electronic Medical Records and Genomics (eMERGE) Phase IV study aims to establish protocols and methodologies for improved genetic risk assessment, and integrate genomic signatures, including monogenic and polygenic risk, and clinical risk factors into routine medical practice to identify individuals at high disease risk and recommend intervention strategies (eMERGE Consortium 2021). Towards this end, the consortium has identified 10 diseases, including T2D, where the predictive power of PRS, combined with clinical risk factors and intervention/treatment paradigms, has shown promise to delay, mitigate, prevent or manage disease. Here, as part of the eMERGE IV study, we present the construction and evaluation of a trans-ancestry T2D PRS to address the opportunities and challenges in the clinical translation of T2D PRS. Specifically, we integrated T2D GWAS in European, African American and East Asian individuals to construct the trans-ancestry PRS using state-of-the-art Bayesian polygenic modeling methods (Ruan et al. 2021), and evaluated the PRS in the multi-ethnic eMERGE I-III samples (Stanaway et al. 2019), four African American cohorts, and the Taiwan Biobank (TWB; Chen et al. 2021). Our efforts represent the first step towards the implementation of T2D PRS into routine clinical care.

## METHODS

### Construction of trans-ancestry T2D PRS from published T2D GWAS

We identified three large-scale T2D GWAS conducted in different populations to derive a trans-ancestry T2D PRS: (i) a GWAS in individuals of European descent with 74,124 T2D cases and 824,006 controls (Mahajan et al. 2018); (ii) a meta-GWAS of T2D in African Americans performed by the MEta-analysis of type 2 Diabetes in African Americans (MEDIA) Consortium with 8,284 cases and 15,543 controls (Ng et al. 2014); and (iii) a GWAS in the Japanese population with 45,383 cases and 132,032 controls, performed by Biobank Japan (BBJ; Sakaue et al. 2021).

We used PRS-CSx, a recently developed Bayesian polygenic modeling method, to construct the trans-ancestry PRS (Ruan et al. 2021). PRS-CSx jointly models the three GWAS summary statistics and couples genetic effects across populations using a shared continuous shrinkage prior, which enables more accurate effect size estimation by sharing information between summary statistics and leveraging linkage disequilibrium (LD) diversity across discovery samples. The shared prior allows for correlated but varying effect size estimates across populations, retaining the flexibility of the modeling framework. In addition, PRS-CSx accounts for population-specific allele frequencies and LD patterns, and inherits efficient and robust posterior inference algorithms from PRS-CS (Ge et al. 2019). We used pre-computed 1000 Genomes Project (1KG; The 1000 Genomes Project Consortium 2015) reference panels that matched the ancestry of each discovery GWAS, and a fully Bayesian algorithm for model fitting, which automatically learned all model parameters from the summary statistics without the need for hyper-parameter tuning. Population-specific posterior effect size estimates were combined using an inverse-variance-weighted meta-analysis (via the “--meta” option provided by the software). The final PRS-CSx output included 1,259,754 HapMap3 variants and their weights, which can be applied to any genotyped individual not included in the discovery GWAS to calculate a polygenic risk score.

### Overview of the evaluation of the trans-ancestry PRS

We first evaluated the trans-ancestry PRS constructed by PRS-CSx among the European, African American and Hispanic/Latino individuals in the eMERGE study (Gottesman et al. 2013; Stanaway et al. 2019; eMERGE Consortium 2021), a consortium of U.S. medical research institutions with a goal to develop and disseminate methods and best practices for utilization of electronic medical records (EMR) in genomic research. To define T2D cases and controls in eMERGE, we validated an EMR-based phenotyping algorithm of T2D to apply across the eMERGE samples (see below). Given that individuals of African descent were underrepresented in the discovery GWAS and thus the predictive performance of the PRS was expected to be lower in African individuals, we next evaluated the trans-ancestry PRS in four independent African American cohorts -- REGARDS (Howard et al. 2005), GenHAT (Arnett et al. 2002), HyperGEN (Williams et al. 2000) and WPC (Limdi et al. 2017) -- collected by the University of Alabama (UAB). In all four UAB cohorts, T2D cases were defined with T2D ICD codes, a single measurement of glucose (fasting glucose ≥126 mg/dL [7 mmol/L] or random glucose ≥ 200 mg/dL [11.1 mmol/L]) or use of any glucose-lowering medications. Lastly, since the number of Asian participants in eMERGE was low, precluding the evaluation of the PRS in the Asian population, we sought to assess the trans-ancestry PRS in the Taiwan Biobank (TWB), a community-based prospective cohort study of the Taiwanese population, aged 30 to 70 years old at recruitment (Chen et al. 2021). TWB participants were interviewed using a structured questionnaire at one of the recruitment centers, which included questions on basic demographic information, lifestyle, environmental exposures, reproductive history, medical history, and family history. Participants with self-reported T2D history were defined as cases in the PRS analysis. After removing 1,776 REGARDS samples that overlapped with the MEDIA study, there was no remaining overlap between the GWAS samples and participants in these evaluation cohorts. Figure 1 summarizes the construction and evaluation of the trans-ancestry T2D PRS in this work. Below we briefly describe the phenotyping algorithm in eMERGE, and the sample characteristics and processing of genetic data in each evaluation dataset.

**Figure 1:**
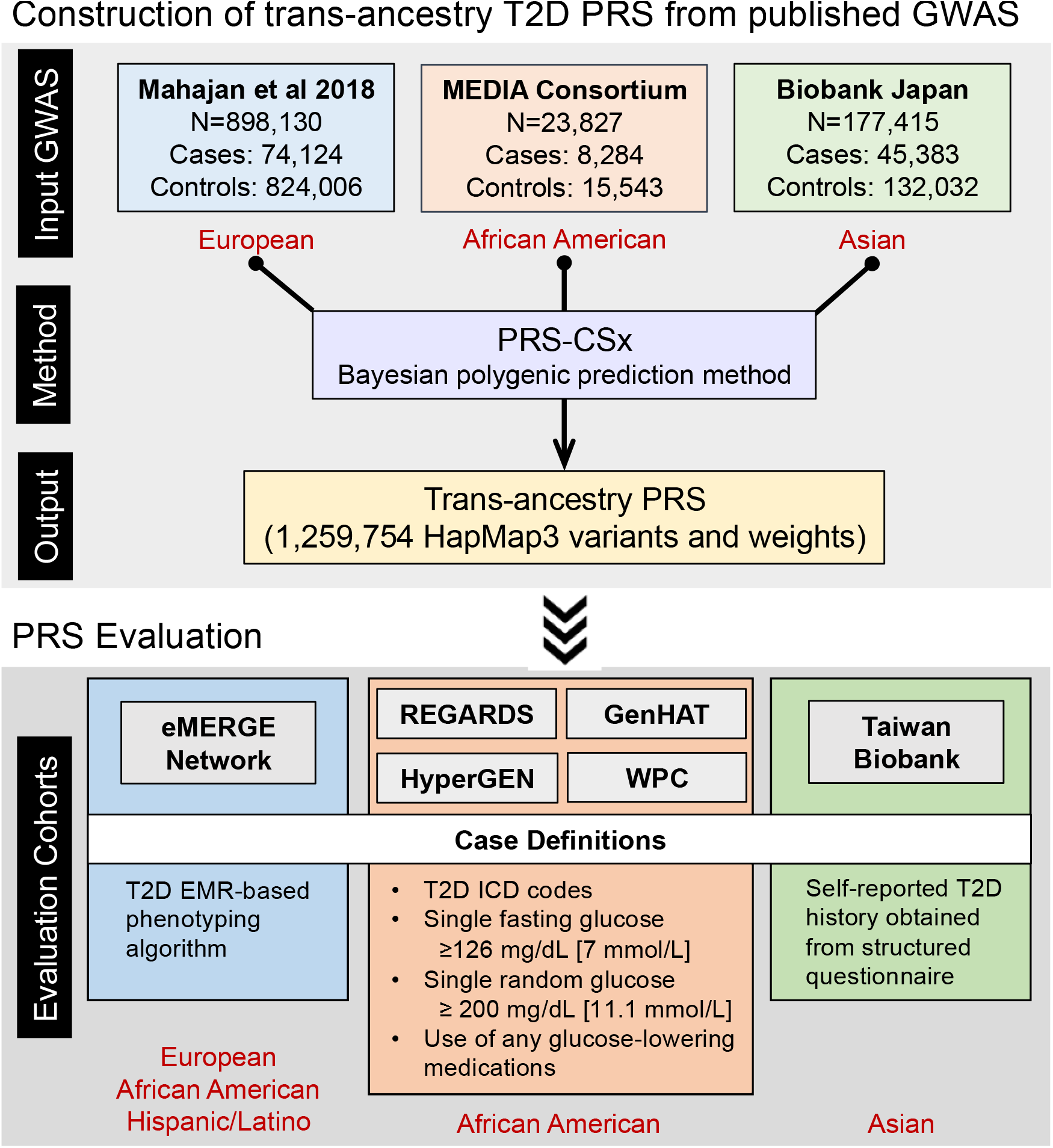
Workflow of the construction and evaluation of the trans-ancestry T2D PRS.

### Validation of EMR-based phenotyping algorithms to classify T2D in eMERGE

We adapted two prior phenotyping algorithms, one from eMERGE (developed in 2011; Kho et al. 2012) and one from Mass General Brigham (MGB) (developed in 2014; Zhang et al. 2019). The eMERGE algorithm was a rule-based algorithm originally developed at Northwestern University. Three eMERGE sites conducted chart reviews of cases and controls defined by the algorithm with sample sizes ranging from 50-100 cases and 44-50 controls, demonstrating 98% positive predictive value (PPV) for cases and 100% negative predictive value (NPV) for controls. The eMERGE algorithm was modified to remove the exclusion of charts with at least one ICD9 code for type 1 diabetes to improve sensitivity, to add ICD10 codes, and to add new diabetes medications released since 2011 (referred to as the “modified eMERGE algorithm”). The MGB algorithm was a machine learning based algorithm using the PheCAP method (Zhang et al. 2019) that had 90% PPV among a chart review dataset that was screen positive for one of 19 phenotypes (prevalence of definite or possible T2D 16%). This algorithm was modified to add ICD10 codes and to add new diabetes medications released since 2014 (referred to as the “modified MGB algorithm”).

We tested the performance of the original eMERGE algorithm, modified eMERGE algorithm, and modified MGB algorithm against two independent gold standard datasets derived from chart reviews, one set from MGB EMR and one set from the UAB EMR. We selected charts for review by applying a data floor of at least one clinical note and at least three ICD9/10 codes from distinct dates, and screened for at least one ICD9/10 code for T2D. We conducted chart reviews for 208 randomly selected screen positive subjects from MGB and 198 screen positive subjects from UAB. In addition, we conducted chart reviews for 200 screen negative subjects from UAB. At MGB, charts were reviewed for diagnostic criteria for T2D by two endocrinology fellows, initially with 20 charts reviewed by both reviewers, to assess concordance. A chart review guideline was developed and used for reviewing the remaining charts, divided into two independent sets. At UAB, charts were reviewed by a trained study coordinator using the same chart review guideline.

Supplementary Table 1 shows the validation results for the three algorithms (eMERGE, modified eMERGE and modified MGB) in the two independent chart review datasets from MGB (138 cases, 70 non-cases; case prevalence 66.3%) and UAB (184 cases, 14 non-cases; case prevalence 92.9%). PPVs ranged from 0.79 to 0.94 with the highest PPV observed for the modified MGB algorithm. Sensitivities ranged from 0.61 to 0.92, with the highest sensitivity observed for the modified eMERGE algorithm. NPVs in the UAB validation dataset were zero for both eMERGE algorithms because all 14 non-cases were classified as cases. Based on these results, we selected the modified MGB algorithm for implementation in eMERGE to define T2D cases and controls for the PRS analysis.

### eMERGE genetic data

We used genetic data from 8 eMERGE sites in this work: Cincinnati Children’s Hospital Medical Center (CCHMC), Children’s Hospital of Pennsylvania (CHOP), Columbia University, Mass General Brigham (MGB), Mayo Clinic, Icahn School of Medicine at Mount Sinai, Northwestern University (NU), and Vanderbilt University Medical Center (VUMC). Imputed genome-wide data against the Haplotype Reference Consortium (HRC; The Haplotype Reference Consortium 2016) across the 8 sites were obtained from the eMERGE Network (Stanaway et al. 2019; eMERGE Consortium 2021). We merged all eMERGE samples with the 1KG phase 3 data (N=2,504), and selected high-quality, common variants shared between the two datasets. We pruned the merged dataset (PLINK command --indep-pairwise 500 50 0.05), retaining a set of independent variants, and calculated principal components (PCs) in the 1KG samples using the LD-pruned variants. We then projected eMERGE samples into the 1KG PC space, and grouped each eMERGE sample with one of the four 1KG super-populations -- European [EUR], African [AFR], Admixed American [AMR], and East Asian [EAS] -- by co-clustering the projected eMERGE samples with the 1KG reference samples. Continental ancestry memberships were verified by visual inspection of the PC plots. We further intersected genetically inferred ancestry with self-reported race/ethnicity, namely White and non-Hispanic/Latino, Black or African American, Hispanic or Latino, and Asian, for the four ancestral groups, respectively, and randomly removed one sample from each pair of related individuals (kingship coefficient >0.1), leaving 54,793 European, 12,472 African American and 2,374 Hispanic/Latino individuals with T2D case and control definitions (Table 1; Supplementary Table 2). We did not use Asian samples in subsequent PRS analyses due to the small sample size. Variants with minor allele frequency (MAF) <1% within each population were excluded.

**Table 1:** Sample characteristics of the evaluation datasets.

| | | Age<br>(Mean ± SD) | Sex<br>(Female %) | Ncase | Ncontrol |
| --- | --- | --- | --- | --- | --- |
| eMERGE | European | 59.4 ± 23.2 | 51.3% | 8389 | 46404 |
| | African | 45.8 ± 22.9 | 60.0% | 2688 | 9784 |
| | Hispanic/Latino | 56.3 ± 20.6 | 60.5% | 868 | 1506 |
| UAB African<br>American<br>Cohorts | REGARDS | 63.8 ± 9.3 | 60.5% | 1659 | 5086 |
| | GenHAT | 66.1 ± 7.5 | 55.3% | 2776 | 2722 |
| | HyperGEN | 47.0 ± 12.8 | 63.5% | 402 | 1494 |
| | WPC | 57.5 ± 15.2 | 57.6% | 300 | 355 |
| Taiwan Biobank | Batch 1 | 48.9 ± 11.1 | 49.3% | 1248 | 23862 |
| | Batch 2 | 50.5 ± 10.5 | 68.6% | 2806 | 51272 |
| | Batch 3 | 49.3 ± 10.9 | 65.7% | 516 | 9862 |

### Reasons for Geographic and Racial Differences in Stroke Study (REGARDS)

REGARDS (Howard et al. 2005) is a national, population-based, longitudinal study of incident stroke and associated risk factors of over 30,000 Black and White adults aged 45 years or older from all 48 contiguous U.S. states and the District of Columbia. REGARDS was designed to investigate reasons underlying the higher rate of stroke mortality among Blacks compared to Whites, as well as why residents of the Southeastern U.S. had worse death rates compared to other U.S. regions. By design, participants were oversampled if they were residents of the stroke belt or if they were Black. Participants completed a computer-assisted telephone interview to collect demographic information and medication adherence, and an in-home visit for blood pressure measurements and collection of blood and urine samples. Participants have been contacted on six-month intervals to obtain information on incident stroke or secondary outcomes. Genotyping was performed on 8,916 Black participants using Illumina MEGA arrays. Imputation was conducted using release 2 (r2) of the National Heart Lung and Blood Institute (NHLBI) TOPMed reference panel available through the BioData Catalyst framework. Participants were excluded if they had sex mismatch or genotyping call rate <0.95, or if they were duplicates or an outlier in the principal component analysis (PCA; outside of 6 standard deviations), resulting in 6,745 individuals (Table 1). Imputed variants were inspected for their imputation quality scores (R^2^) and it was noted that more than 99% of the variants with MAF >1% had an imputation quality >0.6. Given the high-quality in the imputed callset for variants with MAF >1%, genotypes with genotypic probability >0.9 were retained.

### The Genetics of Hypertension Associated Treatments (GenHAT) Study

GenHAT (Arnett et al. 2002) is an ancillary study of the Antihypertensive and Lipid Lowering Treatment to Prevent Heart Attack Trial (ALLHAT). ALLHAT (ALLHAT Collaborative Research Group 2000) was a randomized, double blind, multicenter clinical trial with over 42,000 high-risk individuals who had hypertension, aged 55 years or older, and had at least one additional risk factor for cardiovascular disease (CVD). ALLHAT is the largest antihypertensive treatment trial to date and was ethnically diverse, enrolling over 15,000 Black subjects. Participants were randomized into four groups defined by the class of assigned antihypertensive medication including chlorthalidone, lisinopril, amlodipine and doxazosin at a ratio of 1.7:1:1:1. Due to a significant increase of major CVD outcomes compared to participants on chlorthalidone, doxazosin was discontinued. The original GenHAT study (N=39,114) evaluated the effect of the interaction between candidate hypertensive genetic variants and different antihypertensive treatments on the risk of fatal and non-fatal CVD outcomes. Genotyping was performed using Illumina Infinium Multi-Ethnic AMR/AFR BeadChip (MEGA) arrays on 7,546 Black adults who were hypertensive and randomized to either chlorthalidone or lisinopril. Imputation was performed using version r2 of the NHLBI TOPMed reference panel. Participants were excluded if they failed genotyping, had sex mismatch or genotyping call rate <0.95, or if they were an outlier in the PCA (outside of 6 standard deviations). Since ALLHAT used a fasting glucose ≥ 140 mg/dL for the definition of T2D, we excluded controls that had baseline fasting glucose ≥126 mg/dL (N=201 excluded) or missing a fasting glucose measure altogether (N=1,209 excluded). This resulted in 5,498 individuals eligible for this study (Table 1). Imputed variants were inspected for their imputation quality scores (R^2^) and it was noted that more than 99% of the variants with MAF >1% had an imputation quality >0.6. High-quality genotype calls with genotypic probability >0.9 were retained.

### The Hypertension Genetic Epidemiology Network (HyperGEN)

HyperGEN (Williams et al. 2000) is a cross-sectional, population-based study and component of the NHLBI Family Blood Pressure Program that was designed to identify genetic risk factors for hypertension and target end-organ damage due to hypertension. The cohort is composed of White and Black sibships in which at least two siblings were diagnosed with hypertension (defined as either self-reported use of antihypertensive medications or SBP ≥140 mmHg and/or DBP ≥90 mmHg at two separate evaluations) before age 60, their unmedicated adult offspring, and age-matched controls. Later the study population was expanded to include other siblings of the original sibling pair as well as any offspring for a total sample size of N=5,000. Genotyping on Black participants was performed using whole genome sequencing (WGS), through the NHLBI WGS program (N=1,896; Table 1). In order to harmonize our imputation efforts with the array-based panels of the REGARDS, GenHAT and Warfarin (see below) studies, we compiled a set of non-monomorphic and non-multi-allelic SNPs with MAF >1% that were genotyped as part of those studies. This yielded a total of 2,204,415 SNPs that were used as fence post markers for imputing the HyperGEN cohort using version r2 of the NHLBI TOPMed reference panel. Imputed variants were inspected for their imputation quality scores (R^2^) and it was noted that more than 99% of the variants with MAF >1% had an imputation quality >0.6. High-quality genotype calls with genotypic probability >0.9 were retained.

### Warfarin Pharmacogenomics Cohort (WPC)

The UAB WPC (Limdi et al. 2017) is a prospective cohort of first-time warfarin users aged 19 years or older starting warfarin for anticoagulation. Warfarin therapy requiring a target international normalized ratio (INR) range of 2-3 was initiated in patients with venous thromboembolism, stroke/transient ischemic attacks, atrial fibrillation, myocardial infarction, and/or peripheral arterial disease. Patients requiring a higher intensity (INR 2.5 to 3.5) or lower intensity (INR 1.5 to 2.5) of anticoagulation were excluded. Baseline demographics, as well as medication history and compliance were obtained. Changes in INR, medications and laboratory parameters were documented at each clinical visit as reported previously. Genotyping was performed on the Illumina MEGA array and an Illumina 1M duo array for 599 and 297 Black participants, respectively. Participants aged 40 years or younger were excluded, leaving 655 individuals (Table 1). Imputation was performed using version r2 of the NHLBI TOPMed reference panel. Imputed variants were inspected for their imputation quality scores (R2) and it was noted that more than 99% of the variants with MAF >1% had an imputation quality >0.6. High-quality genotype calls with genotypic probability >0.9 were retained.

### The Taiwan Biobank (TWB)

A total of 110,926 TWB participants were genotyped using two different customized arrays (TWBv1: 27,719 samples; TWBv2: 83,207 samples). Due to data release timelines, samples genotyped on the TWBv2 array were divided into two subsets, with 68,975 samples and 14,232 samples, respectively. Quality control (QC) and imputation were performed on the three batches of data separately. Detailed information on the sample characteristics, collection of phenotypes, and QC procedures can be found in Chen et al. (Chen et al. 2021).

For each batch, we filtered out variants with genotyping call rate <0.98 and samples with call rate <0.98, and removed variants that were duplicated, monogenic or not correctly mapped to a genomic position. We then merged TWB samples with 1KG phase 3 data (N=2,504), and selected high-quality, common variants shared between the two datasets. Next, we performed LD-pruning (PLINK --indep-pairwise 200 100 0.1) and computed PCs of the merged genotype data with LD-pruned variants. Using the population labels of 1KG samples as the reference, we trained a random forest model with top 6 PCs to classify TWB samples into 1KG super-population groups. We retained TWB samples that can be assigned to a homogeneous East Asian group with a predicted probability >0.8. After population assignment, we filtered out outliers in heterozygosity rate and population-specific PCs, and samples with sex mismatch. Imputation was performed using Eagle v2.4 (for pre-phasing; Loh et al. 2016) and Minimac4 (Das et al. 2016) with 1KG phase 3 data as the reference panel. We randomly removed one sample from each related pair of individuals within or across batches, leaving 25,110, 54,078 and 10,378 individuals for the three batches, respectively (Table 1). Post-imputation QC excluded variants with imputation quality scores <0.6 and MAF <0.5%.

### Evaluation of PRS

We assessed the trans-ancestry PRS constructed by PRS-CSx in the European, African American and Hispanic/Latino samples of the eMERGE I-III genotyped dataset, four UAB African American cohorts (i.e., REGARDS, GenHAT, HyperGEN and WPC), and TWB. For each of these evaluation datasets, we calculated the PRS for each individual by multiplying the number of risk alleles by the PRS-CSx inferred weights for each variant and summing across the genome using the --score function in PLINK 1.9 (Chang et al. 2015). To assess the prediction accuracy of the PRS, we calculated the proportion of variation in the T2D case-control status explained by the PRS on the liability scale (Lee et al. 2011), the area under the receiver operating characteristic (ROC) curve (AUC) for the PRS, and the odds ratio (OR) per standard deviation (OR/SD) change in the PRS. We further identified high-risk individuals at the top 2%, 5% or 10% of the PRS distribution, and examined the clinical utility of these classifiers by calculating the sensitivity, specificity, prevalence-adjusted PPV and NPV, and OR of the high-risk individuals versus the rest of the samples. We used population-specific prevalence of diagnosed T2D published recently (European 10.0%; African 12.5%; Hispanic 13.1%; Asian 13.7%; Wang et al. 2021). Prevalence-adjusted PPV was calculated as:

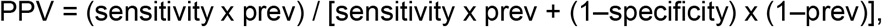

and prevalence-adjusted NPV was calculated as:

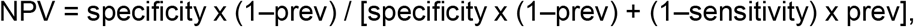

where prev denotes population-specific prevalence. In all the PRS analyses, we adjusted for age, sex and top 10 PCs of the genetic data. For the analysis in the eMERGE study, we additionally adjusted for eMERGE study sites. We obtained an overall OR for the African American population at various cutoffs by meta-analyzing the eMERGE African American dataset with the four UAB African American cohorts using an inverse-variance-weighted approach; we obtained an overall OR for the East Asian population at various cutoffs by meta-analyzing the three TWB batches.

## RESULTS

### Evaluation of the PRS in eMERGE

Table 1 and Supplementary Table 2 show the number of T2D cases and controls in the European, African American and Hispanic/Latino populations, for which we had sufficient sample sizes to evaluate the performance of the PRS, across the 8 eMERGE sites. When the target population was European, the trans-ancestry PRS explained 9.2% of the variation in the T2D status on the liability scale, with an AUC of 0.66 and OR/SD of 1.96 (95% confidence interval [CI] 1.91-2.02), after adjusting for age, sex, top 10 genetic PCs and eMERGE study sites (Table 2; Supplementary Table 3). The PRS provided predictive power above and beyond the covariates, increasing the AUC from 0.74 (the covariates-only model) to 0.79 when combining covariates and the PRS as predictors (Supplementary Table 3). To assess the clinical utility of the trans-ancestry PRS, we compared the risk of T2D among individuals in progressively more extreme cutoffs of the PRS distribution relative to the rest of the samples. Individuals of European ancestry in the top decile of the trans-ancestry PRS had an OR of 3.19 (95% CI: 2.97-3.42; P-value = 1.33E-232) compared with individuals in the bottom 90% of the PRS distribution. Risk continued to increase when contrasting the top 5% of the PRS (OR = 3.55, 95% CI: 3.24-3.90; P-value = 1.88E-158) or the top 2% of the PRS (OR = 4.21, 95% CI: 3.66-4.84; P-value = 1.82E-89) to the lower tails of the PRS distribution (Table 2; Supplementary Table 3). This shows that the trans-ancestry PRS can identify individuals of European ancestry with significantly increased risk of T2D. Using the top 2% of the PRS as the classifier, the prevalence-adjusted PPV and NPV were 0.26 and 0.90, respectively (Table 2; Supplementary Table 3).

**Table 2:** Prediction accuracy of the trans-ancestry T2D PRS in the eMERGE dataset across three populations.

| Population | Liability R2 | Covariate-adjusted AUC | OR per SD (95% CI) | Top 2% PRS |  |  |  |
| --- | --- | --- | --- | --- | --- | --- | --- |
|  |  |  |  | OR (95% CI) | P-value | Adjusted PPV | Adjusted NPV |
| European | 9.2% | 0.66 | 1.96 (1.91, 2.02) | 4.21 (3.66, 4.84) | 1.82E-89 | 0.26 | 0.90 |
| African | 2.8% | 0.58 | 1.54 (1.46, 1.64) | 1.98 (1.43, 2.74) | 4.34E-05 | 0.21 | 0.88 |
| Hispanic | 8.0% | 0.63 | 2.08 (1.84, 2.35) | 6.87 (3.11, 15.15) | 1.81E-06 | 0.43 | 0.87 |

When using the trans-ancestry PRS as a predictor of T2D status for eMERGE African American individuals, the variance explained on the liability scale was 2.8%, the covariate-adjusted AUC was 0.58, and the OR/SD was 1.54 (95% CI: 1.46-1.64; Table 2; Supplementary Table 3). As expected, the prediction accuracy of the PRS in the African American population was substantially lower compared with the prediction in the European population, reflecting the current Eurocentric bias in genomic studies and the fact that our trans-ancestry PRS was constructed from GWAS of predominantly European and East Asian individuals. Nevertheless, the T2D risk increased with higher PRS values and the OR comparing individuals in the top 2% of the PRS distribution with the rest of the individuals was 1.98 (95% CI: 1.43-2.74; P-value = 4.34E-05), indicating that the trans-ancestry PRS can be useful in identifying individuals with elevated T2D risk in the African population (Table 2; Supplementary Table 3).

The prediction accuracy of the trans-ancestry PRS in the Hispanic/Latino group was between that observed in the European and African American individuals: variance explained = 8.0%; AUC = 0.63; OR/SD = 2.08 (Table 2; Supplementary Table 3), reflecting that the Hispanic/Latino population is a recent admixture among Europeans, Africans and Native Americans. Hispanic individuals with high PRS had substantially increased risk of T2D, with an OR of 6.87 (95% CI: 3.11-15.15; P-value = 1.81E-06) when contrasting the top 2% of the PRS with the remaining 98% of the distribution (Table 2; Supplementary Table 3). However, we note that due to the small sample size of the Hispanic/Latino sample in the eMERGE study, the OR estimates for the tails of the PRS distribution were associated with large uncertainties.

### Evaluation of the PRS in UAB African American cohorts

Given that the prediction accuracy of the trans-ancestry PRS in the eMERGE African American samples was relatively low compared with other populations, we performed further evaluation of the PRS in four UAB African American cohorts, namely REGARDS (1,659 T2D cases, 5,086 T2D controls), GenHAT (2,776 cases, 2,722 controls), HyperGEN (402 cases, 1,494 controls) and WPC (300 cases, 355 controls) (Table 1). REGARDS, GenHAT and HyperGEN had largely consistent estimates of performance metrics, and the prediction accuracy was higher relative to the prediction in the eMERGE African American samples. Specifically, the AUC of the PRS, adjusting for age, sex and top 10 genetic PCs, was approximately 0.61, and the OR/SD estimates ranged from 1.70 to 1.85 across the three cohorts (Table 3; Supplementary Table 4). OR estimates contrasting individuals in the tail of the PRS distribution with the rest of the samples increased when more extreme cutoffs were applied; at the top 2% of the PRS distribution, an approximately three-fold increase in T2D risk was observed relative to the remaining 98% of the individuals (REGARDS: OR = 3.04, P-value = 3.87E-10; GenHAT: OR = 2.70, P-value = 8.44E-06; HyperGen: OR = 3.37, P-value = 5.37E-04). The prevalence-adjusted PPVs ranged from 0.26 to 0.34 and all prevalence-adjusted NPVs were around 0.88. WPC had the lowest sample size among the four cohorts and had limited power to assess the tail discrimination of the trans-ancestry PRS, but the OR estimate at the top 2% of the PRS was comparable to the other three cohorts, though not statistically significant (OR = 2.7, 95% CI: 0.80-9.09, P-value = 1.09E-01; Table 3; Supplementary Table 4).

**Table 3:** Prediction accuracy of the trans-ancestry T2D PRS in four African American cohorts.

| Cohort | Liability R2 | Covariate-adjusted AUC | OR per SD (95% CI) | Top 2% PRS |  |  |  |
| --- | --- | --- | --- | --- | --- | --- | --- |
|  |  |  |  | OR (95% CI) | P-value | Adjusted PPV | Adjusted NPV |
| REGARDS | 4.6% | 0.61 | 1.70 (1.58, 1.84) | 3.04 (2.15, 4.31) | 3.87E-10 | 0.30 | 0.88 |
| GenHAT | 3.6% | 0.61 | 1.85 (1.70, 2.01) | 2.70 (1.74, 4.18) | 8.44E-06 | 0.26 | 0.88 |
| HyperGen | 6.2% | 0.62 | 1.75 (1.52, 2.02) | 3.37 (1.69, 6.69) | 5.37E-04 | 0.34 | 0.88 |
| Warfarin | 1.5% | 0.57 | 1.37 (1.13, 1.65) | 2.70 (0.80, 9.09) | 1.09E-01 | 0.07 | 0.87 |

### Evaluation of the PRS in TWB

Since the low sample size of Asian individuals in the eMERGE study precluded PRS analysis, we sought to evaluate our trans-ancestry PRS in the Taiwan Biobank (TWB), in which participants were predominantly Han Chinese. Analysis in the three batches of TWB data (batch 1: 1,248 cases, 23,862 controls; batch 2: 2,806 cases, 51,272 controls; batch 3: 516 cases, 9,862 controls; Table 1) produced highly consistent results. The trans-ancestry PRS was strongly associated with self-reported T2D status, with the variance explained on the liability scale ranged from 12.9% to 15.3%, the covariate-adjusted AUC ranged from 0.68 to 0.70, and the OR/SD ranged from 2.01 to 2.19 (Table 4; Supplementary Table 5). The tail of the PRS was highly discriminative; individuals in the top decile of the PRS had a more than three-fold increase in T2D risk relative to those outside the top decile, and the OR increased to approximately 4.50 when using top 2% of the PRS to identify high-risk individuals (batch 1: OR = 4.62, P-value = 7.47E-31; batch 2: OR = 4.60, P-value = 1.80E-69; batch 3: OR = 4.35, P-value = 1.43E-12). The corresponding prevalence-adjusted PPVs ranged from 0.36 to 0.38, and the prevalence-adjusted NPVs were 0.87 across the three batches (Table 4; Supplementary Table 5). Overall, the trans-ancestry PRS was more predictive in this East Asian sample compared with the eMERGE European samples, likely reflecting the contributions from the BBJ T2D GWAS in the training dataset and the more homogeneous TWB samples relative to the eMERGE study whose sample characteristics may vary across different health care systems.

**Table 4:** Prediction accuracy of the trans-ancestry T2D PRS in the Taiwan Biobank (TWB).

| Batch | Liability R2 | Covariate-adjusted AUC | OR per SD (95% CI) | Top 2% PRS |  |  |  |
| --- | --- | --- | --- | --- | --- | --- | --- |
|  |  |  |  | OR (95% CI) | P-value | Adjusted PPV | Adjusted NPV |
| Batch 1 | 15.1% | 0.70 | 2.19 (2.05, 2.33) | 4.62 (3.56, 5.99) | 7.47E-31 | 0.37 | 0.87 |
| Batch 2 | 12.9% | 0.68 | 2.01 (1.93, 2.10) | 4.60 (3.88, 5.45) | 1.80E-69 | 0.38 | 0.87 |
| Batch 3 | 15.3% | 0.70 | 2.16 (1.96, 2.38) | 4.35 (2.89, 6.53) | 1.43E-12 | 0.36 | 0.87 |

### Meta-analysis

Lastly, we derived overall OR, and prevalence-adjusted PPV and NPV estimates at various percentage cutoffs for the African population by meta-analyzing the eMERGE African American dataset with the four UAB African American cohorts, and overall performance metrics for the East Asian population by meta-analyzing the three TWB batches. Along with the eMERGE European dataset, at least 4,500 T2D cases were available for the three populations, enabling accurate assessment of the tail discrimination of the trans-ancestry PRS. Figure 2 shows that the PRS was highly significantly associated with T2D status across different percentage cutoffs and ancestral groups, with comparable predictive performance in the European and East Asian populations and lower prediction accuracy in the African American population. Individuals in the top 2% of the PRS distribution had significantly increased T2D risk, with the OR estimates ranging from 2.55 in the African American samples to 4.58 in the East Asian samples, suggesting clinical value of the trans-ancestry PRS in diverse populations.

**Figure 2:**
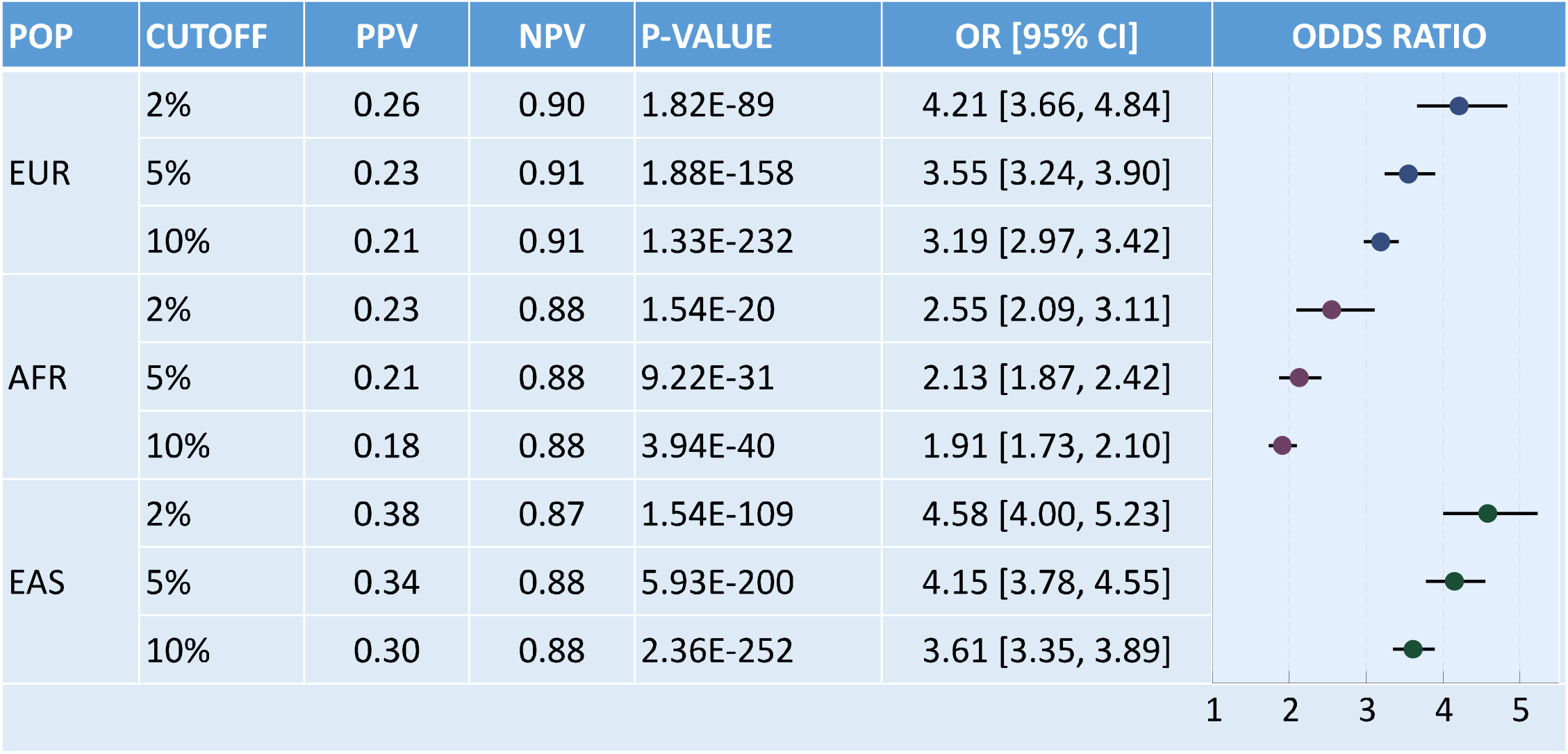
Tail discrimination of the trans-ancestry T2D PRS at various percentage cutoffs in the European, African American (by meta-analyzing the eMERGE African American dataset with the four UAB African American cohorts) and East Asian (by meta-analyzing the three TWB batches) populations. POP = population; EUR = European; AFR = African American; EAS = East Asian; PPV = prevalence-adjusted positive predictive value; NPV = prevalence-adjusted negative predictive value.

## DISCUSSION

We have shown that the top 2% of a trans-ancestry PRS distribution can identify individuals of European, African American, Hispanic/Latino and East Asian ancestry with a roughly 2.5-4.5 fold of increase in T2D risk. By integrating GWAS summary statistics from multiple populations using PRS-CSx, the trans-ancestry PRS was significantly associated with T2D status in all populations examined, providing a robust and potentially clinically meaningful index of risk among diverse patients in clinical settings.

Recent studies have demonstrated in European individuals that T2D PRS can provide predictive power for incident T2D above and beyond established risk factors such as age, body mass index (BMI), smoking, physical activity levels, and history of high glucose and hypertension, and can identify high-risk individuals and stratify lifetime risk trajectories of T2D patients (Läll et al. Genet Med 2017; Lambert et al. Hum Mol Genet 2018), suggesting potential for clinical translation. However, most existing T2D scores were developed and validated in individuals of European descent. As the interest in the clinical implementation of PRS for common diseases like T2D continues to grow, a major challenge is the uncertainty about how best to combine multi-ethnic GWAS and estimate polygenic risk in diverse populations.

In research settings, trans-ancestry PRS are often derived from multi-ethnic meta-GWAS (Mahajan et al. 2020; Polfus et al. 2021) or trained in each target population separately (Marquez-Luna et al. 2017; Weissbrod et al. 2021). However, the former approach does not model population-specific allele frequency and LD patterns, which limits the performance of PRS, while the latter approach requires assigning individuals to discrete ancestral groups to optimize PRS estimation, which is challenging in clinical applications because self-reported race/ethnicity may be a poor proxy for genetic ancestry. The patient group for returning PRS results may also contain admixed individuals who cannot fit into ancestry categories easily. Communication of PRS results in a clinical setting would be facilitated by development of a trans-ancestry PRS without stratifying patients into individual ancestral groups.

In this work, we used a fully Bayesian polygenic modeling method, PRS-CSx, to derive SNP weights for a trans-ancestry PRS. Coupled with post-hoc adjustments that can remove gross differences in PRS distributions across ancestries (see e.g., Khera et al. 2019; Wang et al. 2020), the method can be applied to any target individual without a priori population assignment or hyper-parameter tuning. In addition, PRS-CSx jointly models GWAS summary statistics across populations and explicitly accounts for population-specific LD patterns. While non-European GWAS are often less powerful than European studies, they inform the genetic architecture in non-European populations and may capture population-specific genetic risk factors. Integrating available GWAS across ancestral groups may thus improve the portability of PRS -- especially to non-European populations -- and deliver personalized risk prediction that can more equally benefit all populations. Compared with early T2D PRS developed using a small number of SNPs selected based on statistical and/or biological significance (see Padilla-Martínez et al. 2020 for a review), and more recent T2D PRS derived from large-scale European GWAS (Khera et al. 2018; Mahajan et al. 2018; Vujkovic et al. 2020) or multi-ethnic meta-GWAS (Mahajan et al. 2020; Polfus et al. 2021) using more sophisticated PRS construction methods, our trans-ancestry PRS demonstrated improved prediction accuracy and transferability across ancestral groups, reflecting the combined effect of methodological advances and increased sample sizes and population diversity in the training GWAS.

Our study has several limitations: (i) the gap in prediction accuracy between European/Asian and African populations remains considerable, likely due to the underrepresentation of African samples in the training GWAS; (ii) evaluation samples other than Europeans, African Americans and East Asians were limited; (iii) the sample characteristics of the evaluation cohorts may not fully represent the patient group to which PRS results will be returned, and the effects of PRS may depend on ascertainment and vary across different target samples; (iv) predictive performance of the PRS was only assessed for prevalent T2D cases; and (v) the value of the trans-ancestry PRS over and above standard clinical risk factors remains unclear. Evaluating the capability of the trans-ancestry PRS in identifying incident cases in prospective cohorts and combining common-variant PRS with other genetic and non-genetic risk factors and family history into a Genome Informed Risk Assessment (GIRA), as the eMERGE IV study plans to do, would better define the clinical impact of the PRS.

Previous studies on returning PRS to patients have produced mixed results. For example, the Genetic Counseling/Lifestyle Change (GC/LC) study, which delivered a genetic score constructed from 36 T2D-associated SNPs to overweight patients at risk for T2D, reported limited changes in lifestyles and prevention program adherence compared with controlled participants who did not receive genetic counseling (Grant et al. 2011, 2013; Vassy et al. 2018). The MedSeq Project, which returned polygenic risk estimates for cardiometabolic traits along with monogenic disease risk, pharmacogenomic associations and family history to patients and their healthcare providers (HCPs), found that genetic testing may prompt additional clinical actions of unclear value (Vassy et al. 2017). In contrast, the MI-GENES study provided evidence that disclosure of the genetic risk of coronary heart disease led to greater statin use and lower low-density lipoprotein cholesterol levels after 6 months than returning conventional risk factors alone (Kullo et al. 2016). The different conclusions drawn from these clinical trials may be partly explained by the many factors that influence the effectiveness of returning PRS to patients, including the understanding of the implications of polygenic risk by patients and HCPs, the change of healthcare services based on personalized risk estimates (e.g., surveillance of patients, ordering screening tests, prescribing preventive medications, or providing lifestyle advice), communication of risk to patients, the availability of risk management protocols, and uptake of and adherence to risk-reduction recommendations. Expanded research on the best practice of returning genetic testing results in clinical settings will clarify the benefits of adding polygenic risk estimates to clinical risk factors and family history to create an integrated risk assessment.

In summary, we have constructed and evaluated a trans-ancestry T2D PRS in multiple populations. Future work is needed to expand the scale of non-European genomic resources to further increase the transferability of PRS, assess the predictive performance of the PRS in more diverse samples and real-world settings, refine risk communications, and monitor the impact of returning risk estimates on related clinical outcomes, in order to ensure an equitable and effective deployment of PRS to clinical care.

## Data Availability

GWAS summary statistics from the Mahajan et al. 2018 study are publicly available at the DIAGRAM consortium website (http://diagram-consortium.org). Biobank Japan (BBJ) GWAS summary statistics are publicly available on the BBJ website (https://pheweb.jp). GWAS summary statistics from the Ng et al. 2014 study were obtained through a collaboration with Dr. Ng.
The eMERGE genotype data are deposited in dbGaP (Study Accession: phs001584.v2.p2). Other individual-level datasets analyzed in the present study are not publicly available under current IRB protocols, but are available through authorized collaborations.

## ACKNOWLEDGEMENTS

The eMERGE Network was initiated and funded by National Human Genome Research Institute (NHGRI) through the following grants: U01HG006828 (Cincinnati Children’s Hospital Medical Center and Boston Children’s Hospital); U01HG006830 (Children’s Hospital of Philadelphia); U01HG006389 (Essentia Institute of Rural Health, Marshfield Clinic Research Foundation, and Pennsylvania State University); U01HG006382 (Geisinger Clinic); U01HG006375 (Group Health Cooperative and the University of Washington); U01HG006379 (Mayo Clinic); U01HG006380 (Icahn School of Medicine at Mount Sinai); U01HG006388 (Northwestern University); U01HG006378 (Vanderbilt University Medical Center); and U01HG006385 (Vanderbilt University Medical Center serving as the Coordinating Center). The eMERGE IV Mass General Brigham site was funded by the NHGRI through U01HG008685, the Columbia University site was funded through U01HG008680, and the University of Alabama site was funded through U01HG011167. This work was additionally supported by the following NIH grants: UL1TR001873, OT2OD026553, OT2HL161841, P30AR070253, P30AR069625, R01DK078616 (J.B.M.), R01HL151855 (J.B.M.), R01HL092173 (N.A.L.), R01AR063759 (E.W.K.), R21AR078339 (E.W.K), K25DK128563 (A.K.), K23DK114551 (M.S.U).

